# Inside the black box: Refining intervention theory in the PriDem dementia care study

**DOI:** 10.1101/2025.09.11.25335600

**Authors:** Sarah Griffiths, Emily Spencer, Louise Robinson, Greta Rait, the PriDem Study Team

## Abstract

**Introduction:** The PriDem programme developed a flexible, primary care-led intervention to improve post-diagnostic dementia support, involving Clinical Dementia Leads (CDLs) working with general practices to strengthen care systems. Programme theory was articulated in a logic model, to guide a feasibility implementation study, which demonstrated intervention feasibility, acceptability, and potential for systems-level change. The process of refining programme theory following feasibility testing can appear as a ‘black box’; rarely reported in detail. This paper presents a structured exemplar of theory refinement, addressing this recognised gap in implementation science.

**Methods:** A deductive thematic analysis was conducted, using the logic model as a framework. We synthesised previously reported findings with new qualitative insights from feasibility interviews, fieldnotes, supervision records and researcher reflections. Confirmed, refined, and newly emergent theoretical components were identified and the logic model updated.

**Results:** Many original theory elements were confirmed, including improved review processes leading to enhanced care plan personalisation and staff training increasing confidence in care delivery. New mechanisms were identified, such as mapping local services as a relational tool and care planning templates as educational resources. Pre-implementation activities, such as specific CDL training and champion identification, emerged as critical to success. Role ambiguity and capacity concerns acted as negative mechanisms, impeding implementation. These insights informed a revised logic model to guide future scale-up.

**Conclusions:** This paper demonstrates the value of theory refinement following feasibility testing. By unpacking the ‘black box’ of implementation, we offer a transparent model for optimising complex interventions in primary care-led dementia support.

**Trial registration number:** ISRCTN11677384

## INTRODUCTION

Dementia is a progressive condition impairing memory, communication, and emotional regulation (1). It is estimated that over 900,000 people in England and Wales are affected, with projections reaching 1.7 million by 2040 and care costs rising from £34.7 to £94.1 billion (2, 3). Traditionally led by secondary care, post-diagnostic support is often inadequate, unaffordable, and poorly integrated (4, 5). International policy (6-8) and research (9-12), increasingly advocate for primary care coordination to enhance service access and quality of life for people with dementia (PWD) and their carers (13).

The PriDem research programme (2018-2023) co-developed a primary care-led intervention with PWD, carers, and professionals to improve post-diagnostic dementia care. Adaptable to individual practice needs, the intervention involved Clinical Dementia Leads (CDLs); allied health professionals with dementia expertise, supporting primary care teams to strengthen systems, deliver holistic care, and develop the workforce. Guided by evidence reviews (14-16) and qualitative research (17, 18), a Theory of Change (TOC) outlined how the intervention would drive long-term outcomes (19). The intervention aimed to create systems-level change (20), strengthening local dementia care delivery, with indirect benefits for PWD and carers.

Implementing dementia care interventions is complex, requiring staff time reallocation, workload adjustments, and organisational support (21, 22). A well-developed programme theory supports evaluation by identifying implementation barriers, facilitators, and sustainability strategies (22, 23). To guide implementation, a logic model (S1 File) was developed (28), building on the TOC to address challenges in post-diagnostic care, including inconsistent annual dementia reviews and care planning (24, 25). Following guidance on complex intervention evaluation (23), we then modelled the intervention, conducting a feasibility implementation study to assess change mechanisms and contextual factors (26).

Intervention development best practice involves reflecting on feasibility findings to refine programme theory and guide intervention uptake (22). However, there is limited transparent reporting on this process of reflecting on and iterating programme theory (27), making it difficult to evaluate how and why programmes are adapted over time, and creating a ‘black box’ effect (28, 29). This article exemplifies our approach to this reflection.

### AIMS

- Present an exemplar of using feasibility implementation findings to reflect on intervention programme theory.
- Hypothesise refinements to PriDem intervention theory, to inform a large-scale implementation study.

## METHODS

### Design

A 15-month multi-site, mixed methods feasibility implementation study, was informed by implementation research design tool ImpRes (30) and StaRI reporting standards (31). A ‘Hybrid Effectiveness-Implementation’ design (32) was adopted, whereby the primary aim was to determine the impact of the implementation strategies, and a secondary aim was to assess outcomes. Detailed information on methods has been reported extensively elsewhere (26, 33-35). This article follows Standards for Reporting Qualitative Research (SRQR – see S2 File) (36).

### Ethics

Ethical approval for the study was obtained from Wales REC 4 of the National Research Ethics Service (21/WA/0267). Written or verbal informed consent was obtained for all study participants. Where verbal consent was obtained, this was witnessed and documented in writing by a researcher. The document and audio file were stored on a secure university data management system.

### Setting and participants

Seven general practices were recruited, three in the Southeast and four in the Northeast of England. From practice patient lists and staff, 60 community dwelling PWD, 51 carers and 26 healthcare professionals (HCPs) were recruited between 18^th^ March 2022 and 15^th^ May 2023. Two CDLs, with nursing backgrounds, delivered the intervention over 12 months, one per region, working with practices. They followed an intervention manual, received bespoke PriDem training, clinical supervision from a dementia nurse specialist, and regular sessions with the supervisor and research team.

### Original study overview

The PriDem feasibility implementation study comprised four main elements (Table 1).

**Table 1:** PriDem implementation methods.

| Method | Description |
| --- | --- |
| **Care plan audit** (34):<br>Primary outcome | Adoption of personalised care planning, assessed by comparing the medical records of a random sample of community dwelling PWD pre-intervention and during the 12-months intervention.<br>Audit assessed for presence of a care plan, degree of care plan personalisation and range of care covered. |
| **Questionnaires** (26):<br>Secondary outcomes | PWD and carer quality of life and service use.<br>Data collection: Baseline, 4 months and 9 months.<br>Quantitative outcome measures: DEMQOL (36), EQ- 5D- 5L (37), Neuropsychiatric Inventory (38), Client Services Receipt Inventory (39) to quantify social service use, and medical records to quantify health services use. |
| **Qualitative process evaluation** (35) | A thematic analysis of semi-structured interviews with PWD, carers and HCPs and fieldnotes generated from nonparticipant observations and intervention supervision sessions identified factors influencing implementation. |
| **Feasibility study** (26) | Recruitment and retention rates were examined along with the feasibility and acceptability of the implementation study procedures and the intervention. |

We have previously reported findings from these elements, summarised in S3 File.

### Current study analysis

This paper revisits the programme theory outlined in the initial logic model following feasibility testing. While intended outcomes remain unchanged, the focus here is on the underlying theory, specifically, how outcomes are expected to be achieved. It examines three overlapping intervention strands, each with key targets, activities, implementation strategies (40), and hypothesised mechanisms of change (Table 2).

**Table 2:**
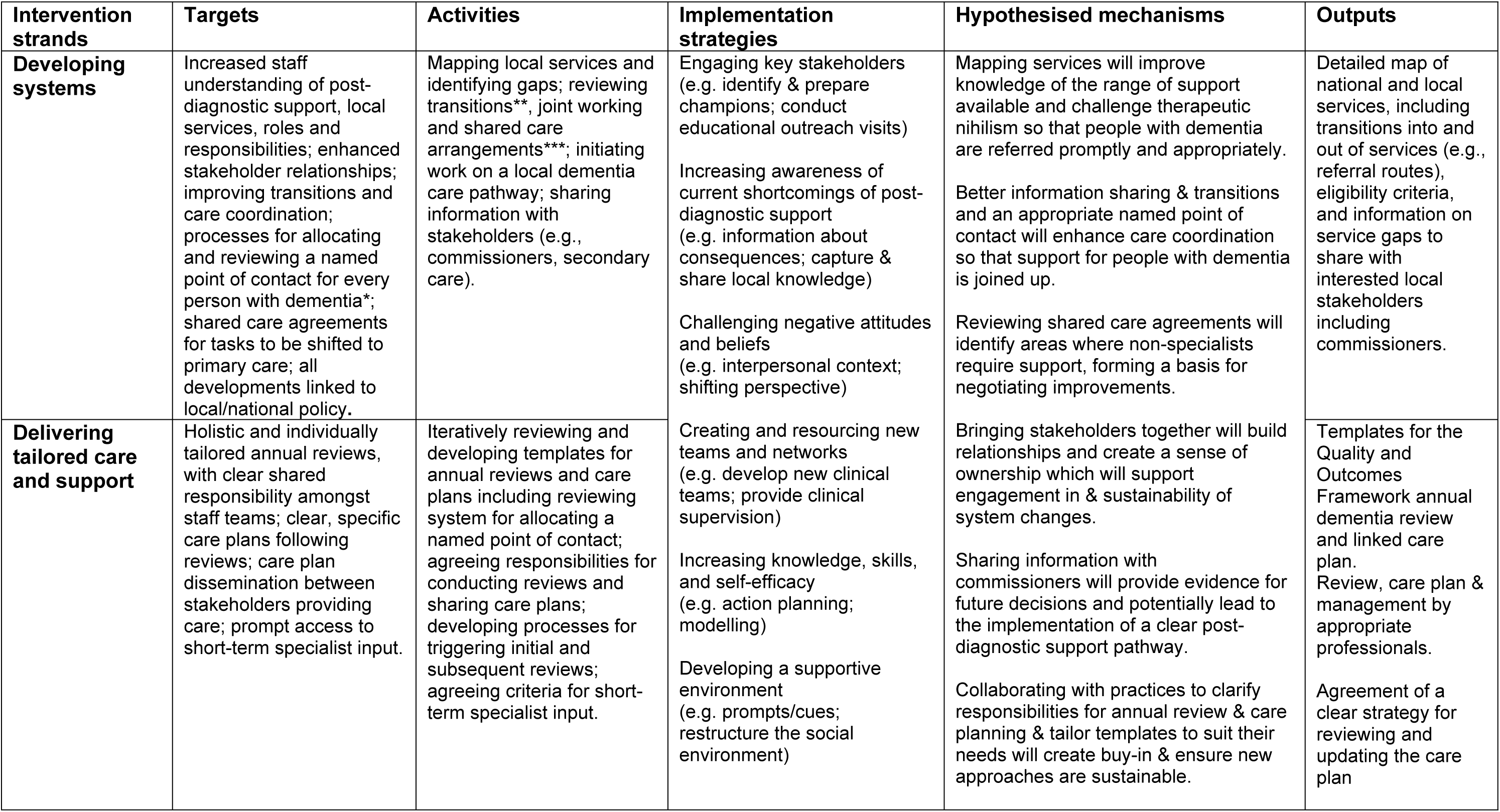
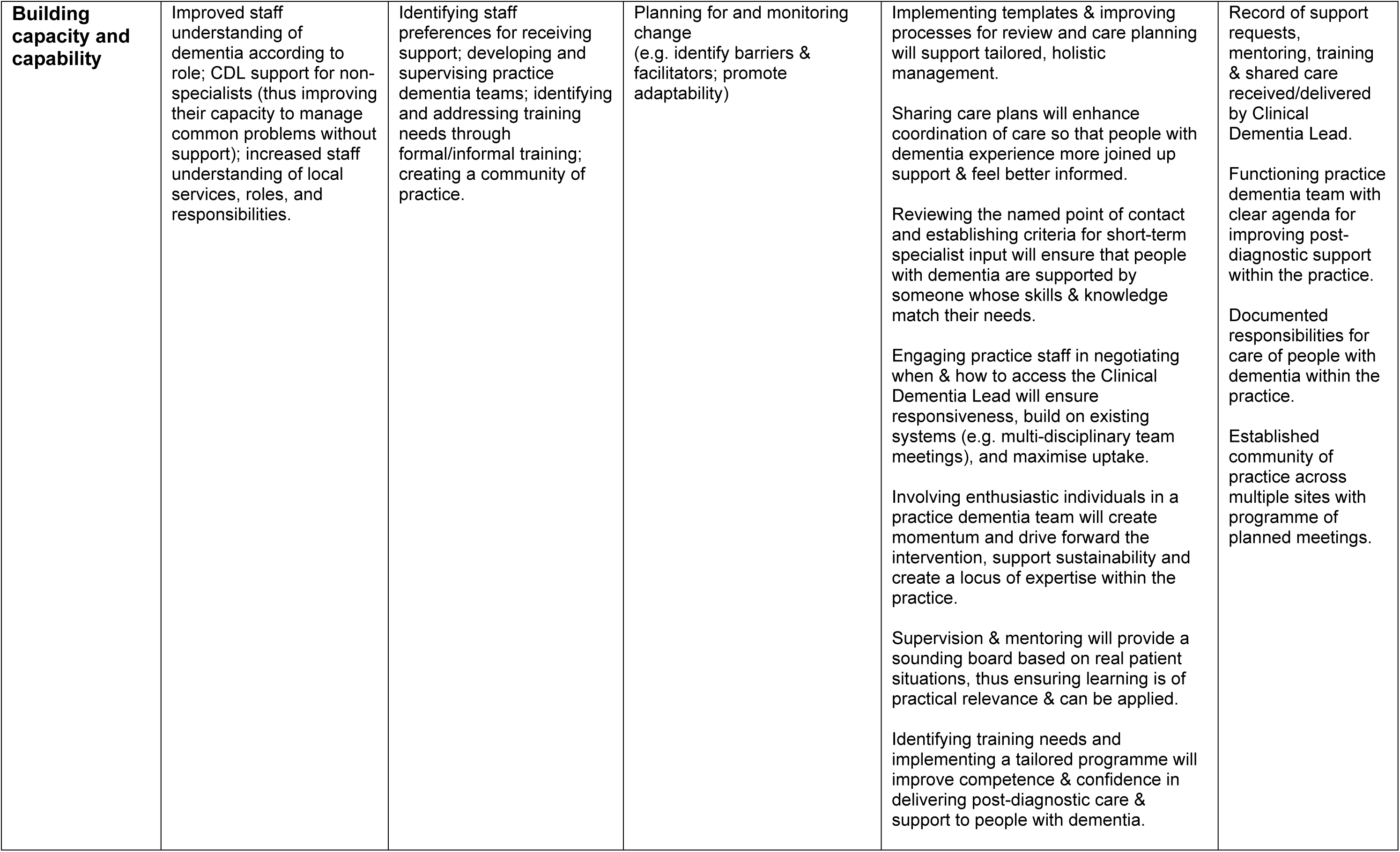
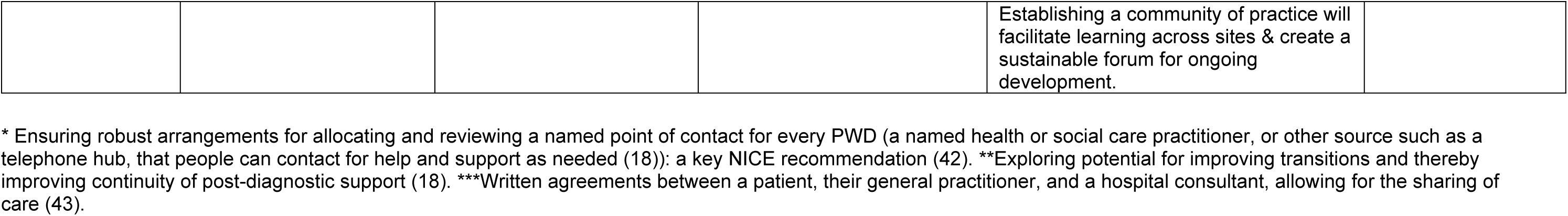
Three PriDem intervention strands, with associated targets, activities, implementation strategies, hypothesised mechanisms and outputs.

We synthesised previously published findings with new insights from feasibility study data, using a fresh analytic lens: exploring how this combined evidence could refine intervention theory in the logic model. Data sources included qualitative interviews, fieldnotes, supervision notes, and researcher reflections, with full details of these sources reported elsewhere (33, 35).

Using Table 2 as a theoretical framework, we conducted a deductive thematic analysis (44, 45), mapping data onto predefined elements. Team discussions then informed refinements to the theory and highlighted untested components. Findings are presented below, with citations for prior work and quotations illustrating new insights.

## FINDINGS

Findings are presented by intervention strand (‘Developing Systems,’ ‘Delivering tailored care and support’ and ‘Building capacity and capability’). Key activities, implementation strategies, mechanisms and intended outputs for each strand are shown in Table 2. We conclude this section by incorporating findings within relevant areas of the original logic model (Table 3).

**Table 3:** Hypothesised Refinements to the PriDem Logic Model.

| Intervention strands | Activities | Implementation strategies | Hypothesised mechanisms |
| --- | --- | --- | --- |
| **Developing systems** | **NEW** Pre-implementation: CDL training and supervision - Understanding the intervention and CDL role within research, legitimising their contribution through resources and real-life examples from the feasibility study, and reflecting on CDL skills and perceived status.<br>✓ Mapping local services and identifying gaps; <u>reviewing transitions, joint working and shared care arrangements;</u><br>✓ initiating work on a local dementia care pathway; sharing information with stakeholders (e.g., commissioners, secondary care).<br>**NEW** Planning early with local stakeholders for:<br>• **Mapping output sustainability; setting realistic timelines.**<br>• **Determining realistic, timed, achievable aims, and acceptable strategies for allocating a named point of contact and implementing shared care agreements.** | Engaging key stakeholders (e.g. identify & prepare champions; conduct educational outreach visits)<br>Increasing awareness of current shortcomings of post-diagnostic support (e.g. information about consequences; capture & share local knowledge). **↻Use of PriDem review and care planning templates as educational tools for this purpose.**<br>Challenging negative attitudes and beliefs (e.g. interpersonal context; shifting perspective)<br>Creating and resourcing new teams and networks (e.g. develop new clinical teams; provide clinical supervision) | **NEW** CDL understanding of the intervention- Insufficient understanding may result in setbacks or stalled progress.<br>**NEW** Mapping as a mechanism as well as an activity - Mapping local services can help build stakeholder relationships and spark collaborative action. <u>Mapping services will improve knowledge of the range of support available and challenge therapeutic nihilism so that people living with dementia are referred promptly and appropriately. Better information sharing & transitions and an appropriate named point of contact will enhance care coordination so that support for people living with dementia is joined up. Reviewing shared care agreements will identify areas where non-specialists require support, forming a basis for negotiating improvements.</u><br>✓ Bringing stakeholders together will build relationships and create a sense of ownership which will support engagement in & sustainability of system changes.<br>✓ Sharing information with commissioners will provide evidence for future decisions and potentially lead to the implementation of a clear post-diagnostic support pathway. <u>Collaborating with practices to clarify responsibilities for annual review & care planning & tailor templates to suit their needs will create buy-in & ensure new approaches are sustainable.</u><br>✓ Implementing templates & improving processes for review and care planning will support tailored, holistic management. <u>Sharing care plans will enhance coordination of care so that people living with dementia experience more joined up support & feel better informed.</u><br><u>Reviewing the named point of contact and establishing criteria for short-term specialist input will ensure that people living with dementia are supported by someone whose skills & knowledge match their needs.</u> |
| **Delivering tailored care and support** | **NEW** Pre-implementation: Study set up should ensure sites understand the intervention and CDL's specialist role in complex dementia cases, not general |  |  |
|  | **caseloads, with agreed definitions of 'complex.'**<br>✓ Iteratively reviewing and developing templates for annual reviews and care plans including reviewing system for <u>allocating a named point of contact; agreeing responsibilities for conducting reviews and sharing care plans; developing processes for triggering initial and subsequent reviews; agreeing criteria for short-term specialist input.</u><br>**NEW** Precursor to the mechanism 'Sharing care plans' - Work with IT specialists, practitioners, and patients to effectively embed documentation in electronic systems. | Increasing knowledge, skills, and self-efficacy (e.g. action planning; modelling)<br>Developing a supportive environment (e.g. prompts/cues; restructure the social environment)<br>Planning for and monitoring change (e.g. identify barriers & facilitators; promote adaptability) | <u>Engaging practice staff in negotiating when & how to access the Clinical Dementia Lead will ensure responsiveness, build on existing systems (e.g. multi-disciplinary team meetings), and maximise uptake.</u><br>↻ **More emphasis and time should be given to the negotiation stage, allowing CDLs, staff, and supervisors to agree on access criteria for specialist dementia input. Discussions should consider MDT hubs and share feasibility insights to support tailored care approaches.**<br>✓ Involving enthusiastic individuals in a practice dementia team will create momentum and drive forward the intervention, support sustainability and create a locus of expertise within the practice.<br>✓ Supervision & mentoring will provide a sounding board based on real patient situations, thus ensuring learning is of practical relevance & can be applied.<br>✓ Identifying training needs and implementing a tailored programme will improve competence & confidence in delivering post-diagnostic care & support to people living with dementia.<br>↻ **Training should include standardised elements around key care planning areas needing improvement, e.g., EoL planning and medication reviews.**<br>↻ **A core element of the training should help staff enhance dementia review practices in line with QoF guidelines and leverage local MDT input.**<br><u>Establishing a community of practice will facilitate learning across sites & create a sustainable forum for ongoing development.</u><br>**NEW** Promoting patient and carer awareness of their right to request annual dementia reviews will lead to improved access to and quality of care. Primary care capacity will have to be carefully negotiated.<br>**NEW** Capacity concerns will lead to a lack of team formation and inaction on improving care planning. |
| **Building capacity and capability** | ✓ Identifying staff preferences for receiving support; developing and supervising practice dementia teams; identifying and addressing training needs through formal/informal training; <u>creating a community of practice.</u><br>**NEW** Pre-implementation: Identifying, training, and supporting champions from the outset and throughout the intervention |  |  |

### 1. Developing systems

This strand addressed gaps in infrastructure supporting primary care-led post-diagnostic dementia care. Despite limited staffing and time, progress was made through stakeholder engagement and identifying service gaps, with findings shared with commissioners to inform local pathway development (35). Insights from this strand also suggest refinements to the intervention theory, as follows.

#### i) New mechanism - CDL understanding of the intervention

An early activity undertaken by CDLs as part of the intervention was to map locally available dementia services. This mapping exercise aimed to identify local post-diagnostic support services for PWD and carers, creating a detailed map for primary care teams and commissioners identifying gaps in provision and informing dementia care pathway development. We have previously highlighted challenges with primary care staff misinterpreting the intervention’s purpose (35). In addition, CDLs initially struggled to comprehend the mapping element, for instance being uncertain whether the purpose was clinical, or research focused. This led to a lack of confidence.

> *‘It kind of made sense as a research nebulous document - but how it would translate clinically into practical tasks…the mapping was really daunting for me. I didn’t really have, I thought, the skills to do it.’* (Interview PROF-01: CDL)

CDLs questioned the value of mapping, believing primary care teams already knew local services and that directories existed. Their uncertainty about their role within a research project further undermined confidence and legitimacy.

> ‘*Why am I…doing something I have no skills and no remit to do, I’ve got no authority. These people are wondering who’s this person who’s a researcher…ringing me to ask me personal stuff about my service, so I don’t have a remit, authority, managerial line for all these organisations.’* (Interview PROF-01: CDL)

In response, CDLs relied on prior clinical experience to interpret tasks. Whilst this can be helpful, it can also hamper learning around what might be both similar and different about the task purpose and required skills.

> *‘I think it’s been quite easy to understand the service mapping because that is something that I’ve done before in other roles….to see what is in the local community and to develop links and relationships with the services. So that’s relatively easy.’* (Interview PROF-04: CDL)

This confusion acted as a negative mechanism, causing early delays in progress. Supporting CDLs to understand the intervention’s goals and their role as interventionists will be essential for successful future implementation.

#### ii) New mechanism - Mapping as a mechanism as well as an activity

Mapping local post-diagnostic support to inform dementia care pathways was a key planned activity. In addition, mapping emerged as a mechanism for building relationships and prompting joint action. For one CDL, mapping enabled connections with working groups, highlighted service gaps, and supported collaboration:

> *‘… it’s putting organisations in contact with each other because they’re all working in silos…saying, look….you’re running an education group for carers, the council already have three years’ experience doing that. Why don’t you talk to each other.*… *mapping turned out to be a good mechanism to engage.*’ (Interview PROF-01: CDL)

The CDL shared finalised service maps with primary care staff and commissioners, highlighting gaps in provision. A condensed ‘desktop directory’ for GPs was also developed. This improved referral routes to dementia advisors and addressed transport issues affecting service transitions and emergency care demand. Mapping also facilitated CDL involvement in local care pathway planning, including commissioner-led sessions.

The approach taken by the second CDL was to develop links with an existing public-facing website, collaborating with providers to improve information accessibility and add information on underrepresented areas e.g., delirium. Primary care practitioners in the region, already reliant on the website for sharing information with patients, steered this approach.

> ‘W*hen [CDL] first started, myself, social prescribers, all different people kept saying, go to [website] for [City]…So [CDL]…worked with them to make sure the dementia information was up to scratch, helped them write some bits…and then brought that back to the GPs*.’ (Interview PROF-13 GP)

However, this community-facing approach risked missing detail on NHS and social care services. Balancing intervention fidelity with stakeholder-driven innovation proved challenging. In this case, the CDL used research evidence, rather than mapping, to influence strategy development:

> *‘The [region] is at a point where they’re just developing their dementia strategy…So I’ve been knocking on doors to try and get into those meetings, to talk about best practice and…bring in…the research behind it.’* (Interview PROF- 04: CDL)

By the intervention’s end, gaps such as poor primary-secondary care communication were identified, but time constraints limited sustained action:

> *‘We’ve looked at…how communication between primary care and secondary care could be influenced…On the back of that there was a…virtual [Multi-disciplinary Team (MDT)] setup with the memory clinic…There has been a pilot to try and address that, but it just hasn’t been utilised.’* (Interview PROF-04: CDL)

Future implementation should ensure CDLs understand the purpose of mapping and its potential to drive collaborative change.

#### iii) New pre-implementation activity - CDL training and supervision

Although we developed and delivered a tailored CDL training and supervision programme, including a practitioner manual, this support for CDLs was not included in our original theory. However, early pre-implementation activities, such as training, are often essential components of a logic model, helping to build readiness, and increase the chances of effective implementation (45). Insights from our feasibility study highlight the need to add pre-implementation activities to our theory, including training and supervision.

As demonstrated earlier in this strand, training and supervision need to help CDLs understand the intervention and their role within a research project and highlight the importance of avoiding assumptions around key activities such as mapping:

> *‘[Supervisor] warns against assuming people know what’s available. Do they refer appropriately, or default to familiar options? Even if staff seem knowledgeable, don’t assume. The map helps ensure timely, informed referrals— practice teams are the key audience. Not all answers lie in existing directories.’* (Intervention supervision notes)

Through intervention supervision, an introductory leaflet for stakeholders was co-developed by CDLs, researchers and supervisors, explaining the project, the CDL role and the mapping, and highlighting benefits for stakeholders. This simple resource helped establish CDL legitimacy and could be included in future training.

Concrete ‘real life’ examples from our feasibility study can be used in future training and supervision to show how the intervention works in practice. Training could also explore CDL skills, perceived status, and roles (e.g., factfinder, influencer) as mechanisms supporting outcomes.

#### iv) New activity - Planning early with local stakeholders for

##### • Mapping output sustainability

The sustainability of service mapping was a concern for both CDLs.

> *‘If I create a directory, it’s as current as it is the day I’ve written it.’* (Interview PROF-04: CDL)

A PriDem mapping template was created and shared with non-study regions (35), demonstrating potential for innovation diffusion (47). By the intervention’s end, a local services map was available on an NHS trust website in one of the regions, with discussions underway to integrate it into general practice systems. However, concerns about ongoing maintenance arose, with social prescribers suggested for oversight. The CDL was unable to support this due to the time-limited nature of the intervention.

Future implementation should prioritise sustainability planning from the outset, involving local stakeholders and setting generous timelines.

##### • Allocating a named point of contact and shared care agreements

There was limited progress on influencing systems for allocating and reviewing a named point of contact for every person with dementia, despite PWD and carers welcoming the idea.

> *‘Well, then you’d know that [practitioner]…whatever the problem was…that would be an advantage I think*.’ (Interview D-07: Person with dementia)

> *‘I just think you might feel a bit more held.’* (Interview C-01: Carer)

CDLs raised the need for named points of contact, but capacity and time constraints hindered system development. Although Dementia advisors and social prescribers were considered for the role, this was not implemented in either region.

The issue of shared care agreements was raised in CDL training:

> *‘[Supervisor] advised shared care agreements tend to be medication focused rather than around other interventions. Key questions will be do people know the shared care agreement exists, resource issues, how concerns/queries are raised when using it.’* (Researcher reflections on CDL training)

However, shared care agreements were not identified as a development priority in either region. Reviewing named point of contact systems and shared care agreements was not achievable within the scope of a 12-month intervention.

For future implementation, early stakeholder consultation could help define realistic, time- bound goals and suitable strategies.

### 2. Delivering tailored care and support

This strand aimed to strengthen post-diagnostic dementia support by addressing gaps in holistic care, standardised review templates, and future planning. CDL engagement and intervention flexibility enabled innovative multidisciplinary approaches, with PriDem templates adapted to local team needs. This led to more personalised care plans and improved holistic care delivery (34, 35). Implementation challenges also informed theory refinement, with key learning points outlined below.

#### i) Refined Implementation strategy - Use of PriDem review and care planning templates as educational tools

Our theory included raising stakeholder awareness of gaps in post-diagnostic support, originally proposed to happen through case note reviews at two time points. Due to staff capacity issues, CDLs struggled to implement this (34). Instead, an adaptable PriDem resource pack with review templates (see S4 File), developed early in the study, proved effective educational tools. CDLs modelled their use by conducting reviews with patients, demonstrating the value of a holistic MDT approach.

> *‘[I said] to the GPs “Oh look, I tried this, and then I could embed the tools into that patient’s notes.” So, it was a like a hook for a GP to say, “I like that review. That was really good.” And then, “Oh but…I couldn’t spend an hour on one patient. Oh but …the receptionist does this bit and the… social prescriber could do that bit…and then maybe I could do my bit.” …So that was…a way for me to say, “Look, this is going to help you because no one’s going to be doing extra work.’* (Interview PROF-01: CDL)

Talking through the template was a vehicle for planning new approaches to team working:

> *‘It’s been really helpful trying to work with [CDL] on the skill mix of… a community team, … [CDL]’s given us the template and tried to help us with the staff mix. And then we’ve looked at what we’ve actually got on the ground and how we can adjust our team to try and accomplish that template as best we can.*’ (Interview PROF-18: GP)

A future theory iteration should incorporate use of PriDem review and care planning templates as educational tools within the range of available implementation strategies.

#### ii) Refined mechanism – Engaging practice staff in negotiating when and how to access the CDL

CDLs were expected to hold a small caseload of complex dementia cases, offering short-term specialist input and modelling good practice. This was to be guided by negotiated criteria with practice staff (Table 2). In reality, defining the CDL clinical role proved complex, with little evidence of effective negotiation. Some staff viewed CDLs as an extra resource for managing the full dementia caseload, rather than as agents of systemic change (35). One CDL addressed this by directing referrals to an existing MDT, with a different but related specialty to dementia, highlighting its governance and educational value.

> *‘When I let the surgeries have too much reign, they just sent me really scrappy emails with half a bit of information about a patient and expected me alone with no back up, no infrastructure, no way patients could contact me…I didn’t think it was safe. I utilised the [specialist MDT] infrastructure and said, can you refer via [them]?…it’s a good place for me to…work out what their needs are then take that back to the [specialist MDT] and demonstrate the PriDem [approach]… and all the GPs are sitting there. So, then they take that learning for the next patient they see.’* (Interview PROF-01: CDL)

This aligns with our mechanism of negotiating referral criteria, which incorporates building on existing systems, however, the approach lacked flexibility when there was an urgent need, and did not allow for proactive, timely, and tailored input.

> *‘One of the GPs raised a patient she had seen recently with complex needs, who had bitten her husband while in the surgery waiting room. There was an obvious need for additional support, GP hadn’t been sure whether to contact safeguarding… general consensus of urgency…CDL suggested referring the patient to the [specialist MDT]…for a meeting in one week.* **Researcher reflection:** *This seems like a missed opportunity…a patient who could use quick input with a dementia-specific focus…This seems the exact sort of complex case that we envisaged the CDL taking on.’ (Observation of practice meeting)*

Lack of negotiation in this case led to both the CDL and practice staff feeling unsupported.

CDLs often expressed uncertainty in supervision, around the amount of direct patient contact expected as part of their roles and the definition of a complex case. Whilst not formally defined, this was described by the supervisor as follows:

> *‘It would be like be when there is perhaps lots of co-morbid conditions and there’s perhaps challenging relationships, or there’s a lack of engagement.’* (Interview PROF-15: CDL Supervisor)

To address these issues, more emphasis and time could be given to the negotiation stage, where CDLs, practice staff and supervisors agree processes and criteria for accessing specialist dementia input, in a framework within which CDLs also feel supported by the practices and satisfied in their work.

Contextual infrastructure such as existing MDTs could be discussed, in terms of whether and how they can support specialist dementia input. As part of our study, some practices developed innovative approaches to delivering tailored care and support, such as One Stop Shop MDT dementia review clinics (35, 48). Future implementation can include sharing these innovations with CDLs and practice staff as illustrative case studies, thereby supporting care delivery decision-making.

#### iii) New pre-implementation activity - Study set up

Effective negotiation depends on a clear understanding of the intervention’s purpose, which some practice staff lacked (35). Future implementation studies should build in more time for educating practice staff and CDLs about the intervention’s nature and the CDL role, fostering a shared understanding. These activities should be included in pre-implementation activities.

#### iv) Refined mechanism - Identifying training needs and implementing a tailored programme: Targeting specific areas of care

Although the feasibility study showed improvements in care planning (26), some areas of care saw less change in representation within care plans, such as medication reviews, and end of life (EoL) planning (34). Given the need to minimise polypharmacy (49) and the challenges of timing EoL discussions (49) future iterations of the intervention could target these specific areas through the mechanism of CDL-led staff training.

#### v) New mechanism – Promoting patient and carer awareness

PWD and carers are typically unaware of their entitlement to an annual review (34). While their role in care delivery was not part of our intervention theory, raising patient awareness of their rights could be a powerful mechanism for improved care, empowering patients and carers to request holistic reviews. This would require careful negotiation of primary care capacity to support increased uptake and provision. We saw evidence of buy-in from some staff, who anticipated that spending more time improving review quality would offer long-term efficiency gains, by enhancing anticipatory care (48).

#### vi) New activity – Precursor to the mechanism ‘Sharing care plans’

Our logic model proposes that sharing care plans between practitioners, PWD and carers enhances care coordination. However, this rarely occurred, largely due to the lack of integrated electronic record systems (34). The PriDem care plan template was not shared with patients as was intended, with practitioners reporting that recording care plans on IT systems as well as creating patient-friendly versions was too burdensome:

> *‘The [PriDem care plan template]…we didn’t really use…it’s a really nice concept, but not logistically achievable to document on your own computer system, which is your Medico-Legal note…and then…create a written care plan…but it might be interesting to see is there a way of doing an [electronic] template that…could be printed out and posted afterwards to the patient…if [it] had the kind of trimmings…to make it accessible… and that was completed as you were going through your review… would definitely be an incredibly valuable addition.’* (Interview PROF-06: GP)

Intervention theory should include activities to support care plan sharing, involving collaboration with primary care IT specialists, practitioners, and patients to embed documentation in electronic systems effectively.

### 3. Building capacity and capability

This strand tackled key challenges in primary care dementia support, such as limited training, inconsistent specialist input, and low staff confidence. In several practices, CDLs identified and engaged motivated staff, from care coordinators to GPs, who championed dementia care and fostered a sense of intervention ownership (35). This enabled tailored training, including joint visits and case discussions, leading to improved staff confidence and competence in post-diagnostic care (35). We identified the following theory refinements.

#### i) New pre-implementation activity – Identifying, training, and supporting champions

Champion identification was a key implementation strategy, with champions proving instrumental in driving meaningful change and supporting programme theory. However, not all practices were able to identify champions, highlighting the need for early identification and sustained support during pre-implementation and beyond. In one practice, running One Stop Shop MDT clinics helped sustain a dementia team:

> *‘Running the one stop shops has probably been one of the key ways of maintaining that… there’s a kind of focus for the team to come together… building an interest and upskilling going forward.’* (Interview PROF-06: GP)

This team continued beyond the study, independent of CDL input, driven by committed champions. Strong leadership and a shared purpose, beyond simply improving dementia care, helped the team thrive. This aligns with the need for ‘clear agendas’ (Outputs, Table 1).

#### ii) New Mechanism – Capacity concerns limiting team formation and care planning

While our theory acknowledges capacity as a contextual factor, new insights suggest it also acts as a mechanism that can hinder team formation and care planning. Although the PriDem approach aimed to optimise existing resources (26), limited staff capacity and lack of funding were major concerns in some practices.

CDLs often engaged champions informally, but staff were reluctant to create formal teams due to workload concerns.

> *‘The perception is that this would be a big task… it would probably need some sort of resourcing to make it attractive.’ (Interview PROF-02: GP)*

One CDL noted that calling groups ‘dementia teams’ was counterproductive, as it implied extra work. Instead, they engaged individuals based on enthusiasm and availability. This may explain the discrepancy between the CDL reporting the existence of operating practice dementia teams and the fact that those identified as team members did not perceive themselves as being part of such a team, suggesting varied perceptions of team membership.

Capacity concerns also affected care planning. Despite progress in care planning, time constraints limited holistic reviews. Many practitioners were unaware that the Quality and Outcomes Framework (QoF) guidelines (51), an annual incentive program for GP practices in England, proposed up to 30 minutes for dementia reviews.

> *‘How do you integrate that into a normal 10-minute appointment?’* (Interview PROF-13: GP)
>
> *‘They’ve got 10-minute appointments, got to try and cram in as much as they can and for a dementia review, it takes much longer.’* (Interview PROF-20: Social Prescriber)

These challenges suggest that without adequate time, team structures and awareness of guidelines, long-term sustainability and quality care planning are at risk.

#### iii) Refined mechanism - Identifying training needs and implementing a tailored programme: Training should help staff enhance dementia review practices in line with QoF guidelines and leverage local MDT input

To mitigate challenges outlined in this strand, we propose that CDL-led staff training and awareness initiatives should go beyond skill-building to include strategies for refining dementia review approaches based on current QoF guidelines, MDT support, and leadership-driven team cohesion.

Table 3 integrates findings into the logic model, indicating which theoretical elements were supported, newly proposed or refined, and which remain untested.

## DISCUSSION

We have presented a detailed reflection on the feasibility implementation of the PriDem intervention, offering critical insights for refining programme theory and informing future large-scale implementation. Findings highlight challenges of translating theory into practice, including role clarity, training, and system integration. This supports guidance emphasising iterative theory refinement in developing complex interventions (23).

### Theory components confirmed, refined, and newly identified

Many original theory elements were supported. For example, enthusiastic individuals drove momentum, templates and improved review processes enhanced care plan personalisation, and staff training increased confidence and competence in care delivery.

In addition, we identified new theoretical components and refined existing ones. Mapping emerged not only as a planned activity but also as a mechanism for interdisciplinary collaboration and service improvement; an intervention diffusion effect (47, 52). Templates, beyond standardising holistic care, functioned as educational tools to challenge therapeutic nihilism and support team-based planning. This contributes to primary care knowledge mobilisation literature (53) by highlighting the need to tailor resources in ways that feel relevant and acceptable to users.

Our refined programme theory includes negative mechanisms; factors that actively hindered implementation. For instance, CDL uncertainty about mapping delayed progress, while unclear roles and perceived lack of role legitimacy limited early stakeholder engagement. These mechanisms, absent from the original theory, reveal active processes disrupting outcomes, thereby strengthening the logic model’s explanatory power for future implementation (54).

Capacity constraints and organisational readiness were influential contextual factors, but our analysis suggests these also function as mechanisms, shaping staff engagement, care planning practices, and sustainability potential. This highlights the need for implementation theory to incorporate not just resource realities but human behaviour in response to them (55). Future studies could draw on change resistance literature (56) and examine the contextual and cultural factors influencing why some practices innovate more than others, including related behaviour change strategies (57).

The role of CDLs in negotiating their clinical input emerged as an area needing refinement. While the logic model assumed joint planning of referral processes, this proved challenging. There is a need for clearer role negotiation processes, underpinned by supportive structures such as existing MDTs and primary care digital systems. Role clarity and decision-space (practitioner autonomy to make decisions about patient care) have previously been identified as key determinants of implementation success (58, 59) with lack of clarity a key source of team friction.

### Implications for future implementation

Our findings advocate for the inclusion of pre-implementation activities in programme theory (46), including CDL training and supervision, study set up activities focused on ensuring understanding of the intervention, and early champion identification. Individuals with a commitment to dementia care and strong leadership qualities, such as persuasiveness and collaboration, can help overcome resistance, break down silos, and foster implementation success when supported from the outset (60). We have previously highlighted the importance of embedding CDLs in practices through physical space and visible presence

(26). Strategies to achieve this should be part of pre-implementation negotiations with practices. Sustained engagement of stakeholders to jointly set realistic goals and timescales is essential for future success of implementing aspects such as allocating named points of contact and sharing care plans. Engagement of stakeholders responsible for IT systems and workforce development is important for supporting activities such as care plan sharing and MDT-based reviews.

The revised logic model (Table 3) identifies aspects yet untested, such as strategies for shared care agreements and establishing a community of practice. This will help guide future inquiry and consequent theory iteration. Given the variable success of dementia practice teams, future implementation might consider dementia support teams across a wider area spanning primary, social and secondary care within a locality. This could prove a powerful mechanism for improving those aspects yet untested, as well as supporting the sharing of care plans between services.

Although our original theory did not account for the role of patients or carers in service delivery, given the systems-level approach, through conversations with research participants, we began to see the potential in empowering people with dementia and their carers to actively request a holistic dementia review and care plan. This led us to hypothesise that such an approach could help improve access to and quality of care. It also opens the possibility of evaluating the impact of a patient awareness-raising mechanism.

Ahead of future implementation, it will be essential to incorporate current contextual factors and national guidelines, particularly those outlined in the government’s 10-Year Dementia Plan (8) and the wider Health Plan for England (61), which both highlight a strategic shift toward community-based, integrated care models to better support people with dementia.

### Strengths and Limitations

We address a recognised gap in implementation science, where theoretical assumptions are often underreported or insufficiently tested, contributing to the ‘black box’ problem in complex interventions (28, 29). Our synthesis draws on multiple data sources - qualitative interviews, fieldnotes, supervision notes, and researcher reflections - viewed through a logic model lens. This strengthens the revised programme theory and supports its potential transferability to other primary care-based dementia interventions.

While no new empirical data are presented, the article integrates feasibility findings to reflect on theory development. As such, insights are context and time specific, and some original theoretical components remain untested and speculative, flagged for future research.

Despite these limitations, the paper offers a strong example of iterative theory development, promoting greater transparency and reproducibility in designing and implementing complex health interventions.

## CONCLUSION

This study demonstrates the value of systematically reflecting on and iterating programme theory in complex intervention research. Our analysis bridges the ‘black box’ of intervention delivery and theory refinement, offering a transparent model for others seeking to optimise dementia care or similarly complex interventions. By adapting theory to practice-informed insights, we enhance the likelihood of effective, scalable, and sustainable implementation in primary care-led dementia support.

## Data Availability

Quantitative data are available from the authors upon reasonable request with permission of Professor Greta Rait. Requests to use data will be submitted on a standard form and reviewed by a committee prior to data-sharing agreements being developed. Ethical approval for this study, as provided by Wales REC 4 of the National Research Ethics Service (21/WA/0267), prohibits us from sharing the full data. Pseudonymised excerpts of data relevant to the analysis have been provided within the paper. While ethical approval is in place to share excerpts of pseudonymised data within published works, the nature of the data mean that transcripts being made available in their entirety could risk participants becoming identifiable. Data being made available in this way would also violate the agreement to which participants consented. Queries relating to data access can be addressed to the Clinical Trials Unit of the UCL Research Department of Primary Care and Population Health via.

## Acknowledgements

We would like to thank: Claire Bamford, Alison Wheatley, Greta Brunskill, Marie Poole, Katie Flanagan, Jane Wilcock, Kate Walters and all those involved in delivering the PriDem intervention.

## SUPPORTING INFORMATION

**S1 File – PriDem Logic Model**

**S2 File - Standards for Reporting Qualitative Research (SRQR) Checklist**

**S3 File - Summary of evaluation findings already shared**

**S4 File - Adaptable PriDem resource pack with review templates**

## Notes

### Competing Interest Statement

The authors have declared no competing interest.

### Clinical Trial

ISRCTN11677384

### Funding Statement

Yes

### Author Declarations

Wales REC 4 of the National Research Ethics Service (21/WA/0267)

